## Supplementary material for "Deep representation learning for clustering longitudinal survival data from electronic health records": SOM file

### Supplementary Note 1

First, for each ICD-10 code, its subcategory (e.g. K50.1), category (e.g. K50) and block (e.g. K50-K52) were extracted using the algorithm below.

**Algorithm ICD-10 mapping**

```
# x is the input ICD-10 code
function ICD-10_map (x):
    if x is in block level:
        return (x, x, x)
    else if x is in category level:
        # block(x) returns the block of the ICD-10 code from a lookup table
        return (x, x, block(x))
    else:
        # category(x) returns the block of the ICD-10 code from a lookup table
        return (x, category(x), block(x))
```

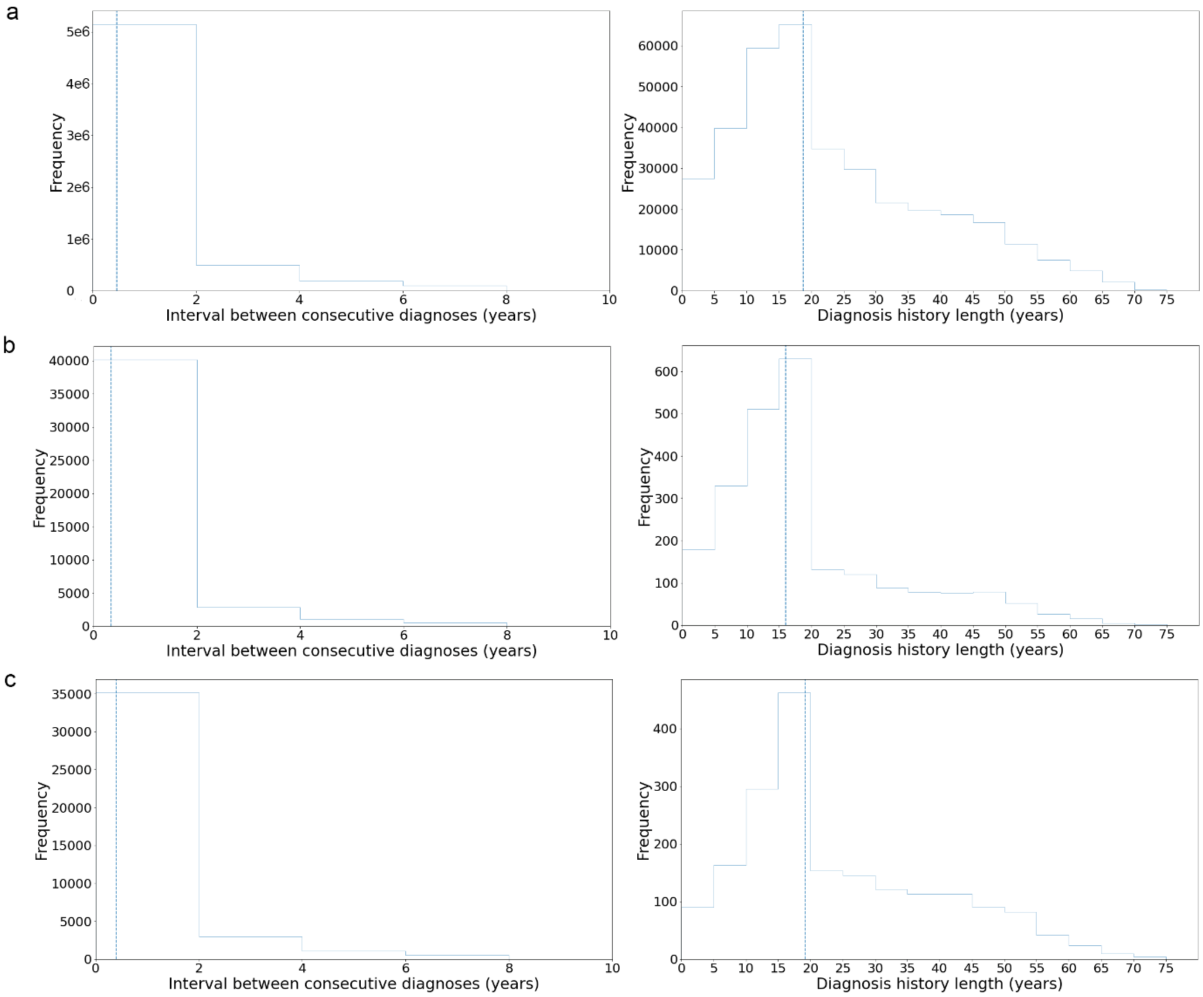


**Supplementary Fig. 1: Summary of EHR data (supplementing Table 1).** Interval between consecutive visits (left) and diagnosis history length (right) for **a** Overall UKBB EHR (Table 1, column 3), **b** T1D/T2D benchmark (Table 1, column 4), and **c** CD use case (Table 1, column 5).

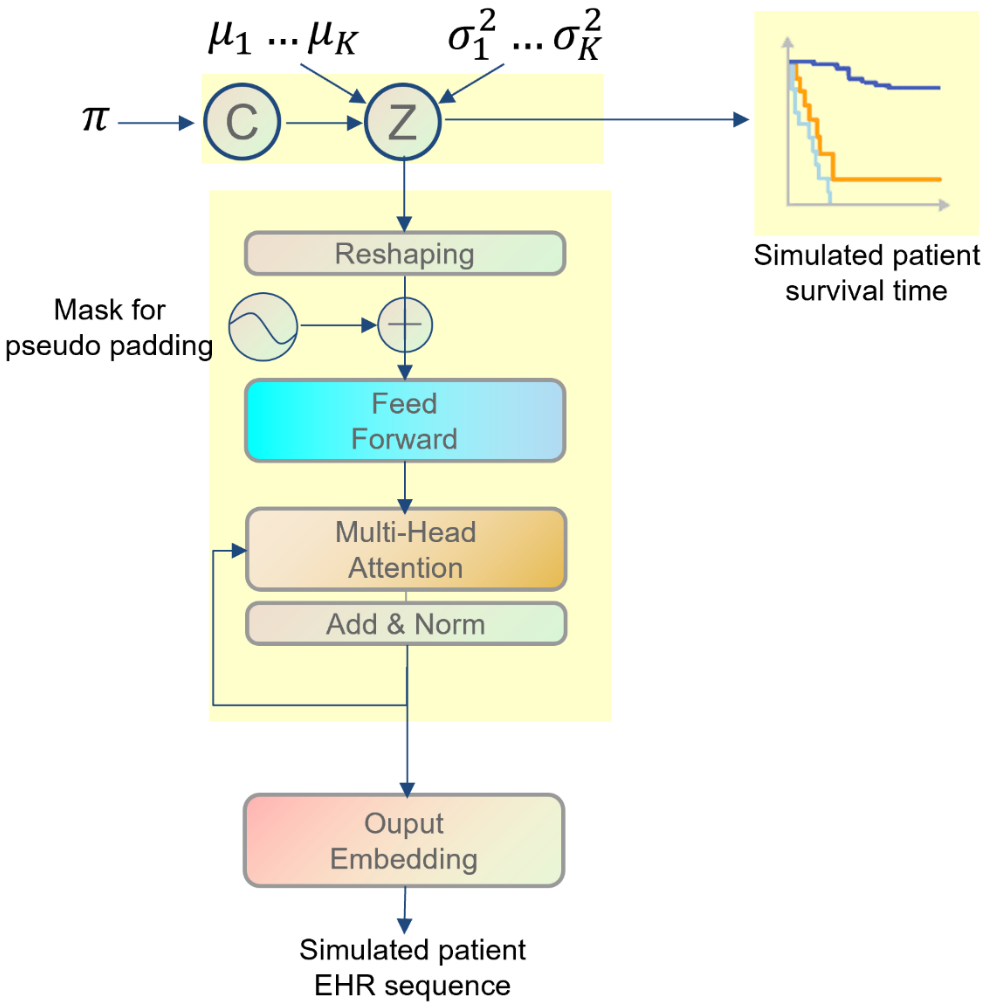


**Supplementary Fig. 2: The generative process of VaDeSCEHR.** For randomly generating one EHR sequence with corresponding time-to-event, we first sample a vector Z from a Gaussian mixture distribution initialized with random parameters. Z is then used to generate both a time-to-event and an EHR sequence. The time-to-event is generated by sampling from a $Weibull$ distribution. The EHR sequence is generated by feeding Z into a transformer-based decoder with randomly initialized weights and then converted to an ICD-10 code using a softmax function. For details, we refer the reader to the Methods section.

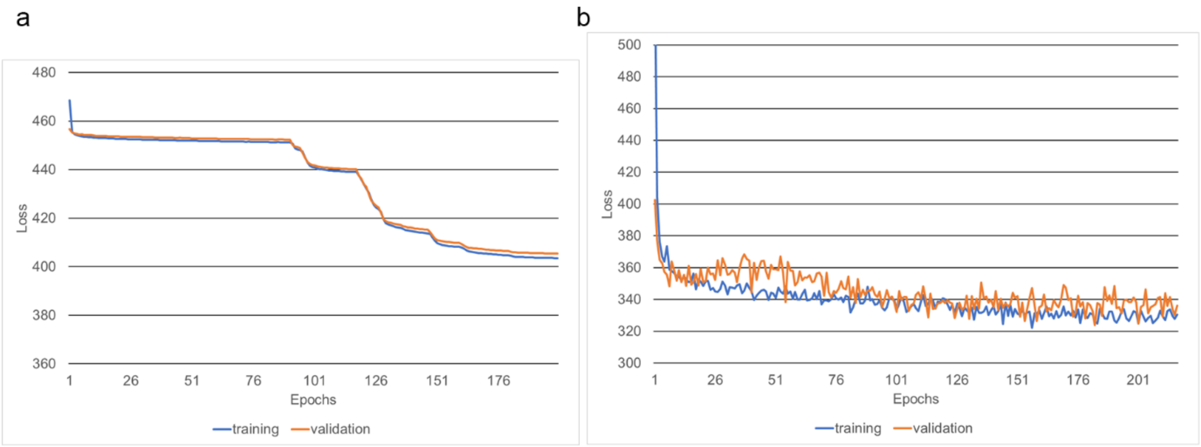


**Supplementary Fig. 3: Overall loss for benchmark analysis. a** synthetic benchmark. **b** T1D/T2D benchmark.

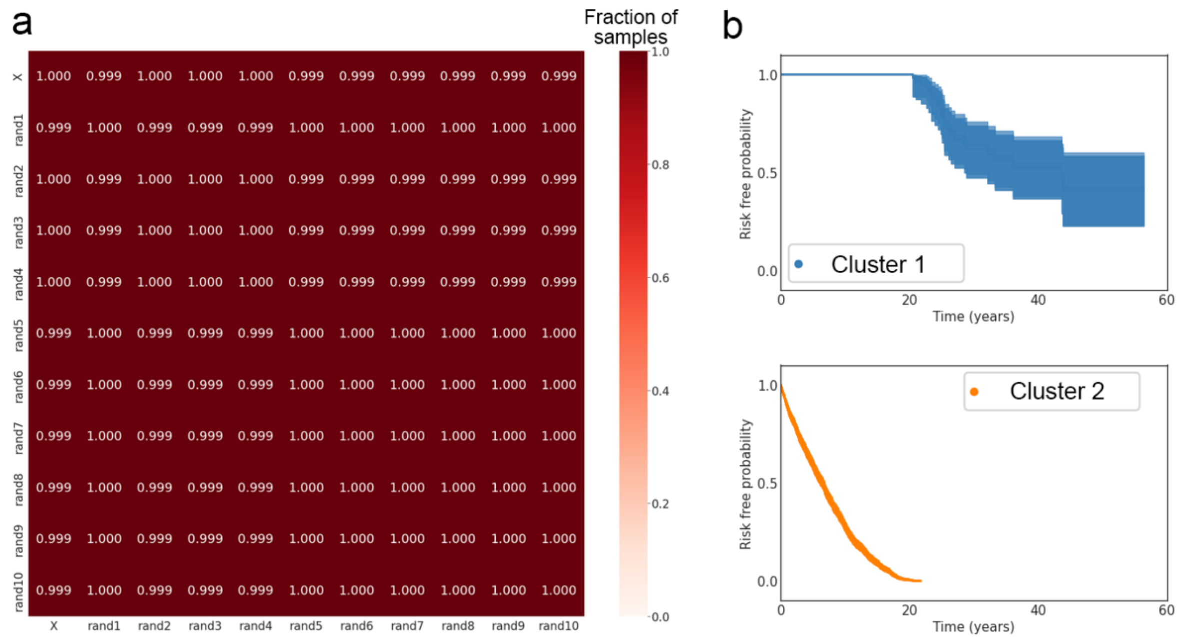


**Supplementary Fig. 4: Stability analysis for T1D/T2D benchmark.** Results of running the VaDeSCEHR 10 times with different initial embeddings and weight initializations (rand1-10, n =10). X indicates the result that we report as the main result in the manuscript. **a** Stability analysis for clustering. Each value indicates the fraction of the samples that obtain the same cluster assignment between two models. **b** Stability analysis for risk modeling. For each cluster, we plot the survival curve from 10 different models. Survival curves show a high degree of similarity across runs.

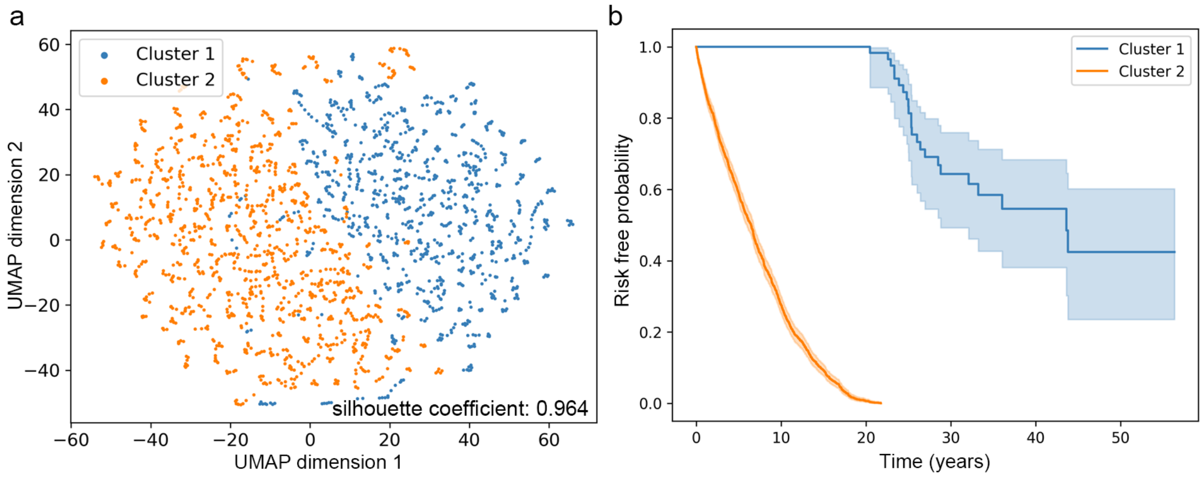


**Supplementary Fig. 5: Using VaDeSCEHR to distinguish T1D from T2D patients. a** UMAP of the patient representations (silhouette coefficient: 0.964). **b** Cluster-specific Kaplan–Meier curves for both clusters.

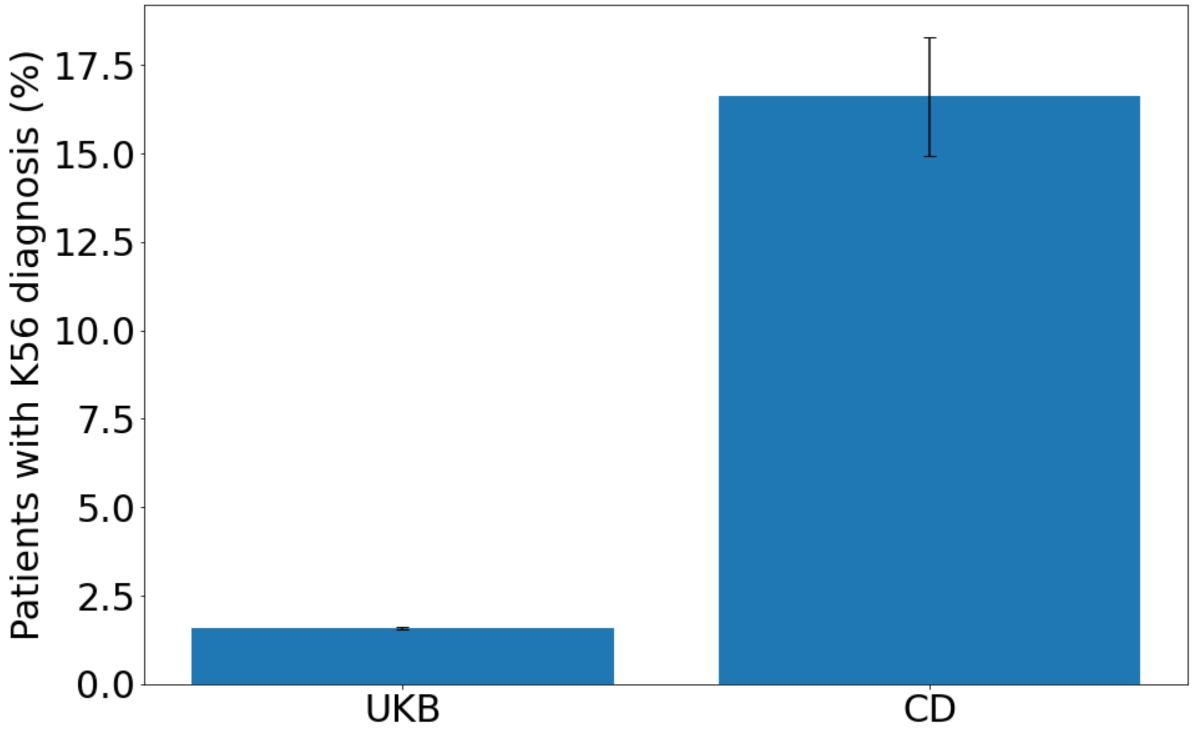


**Supplementary Fig. 6: Enrichment of K56 in CD patients compared to UK Biobank patient population.** Two sided Chi-squared p-value < 0.0001.

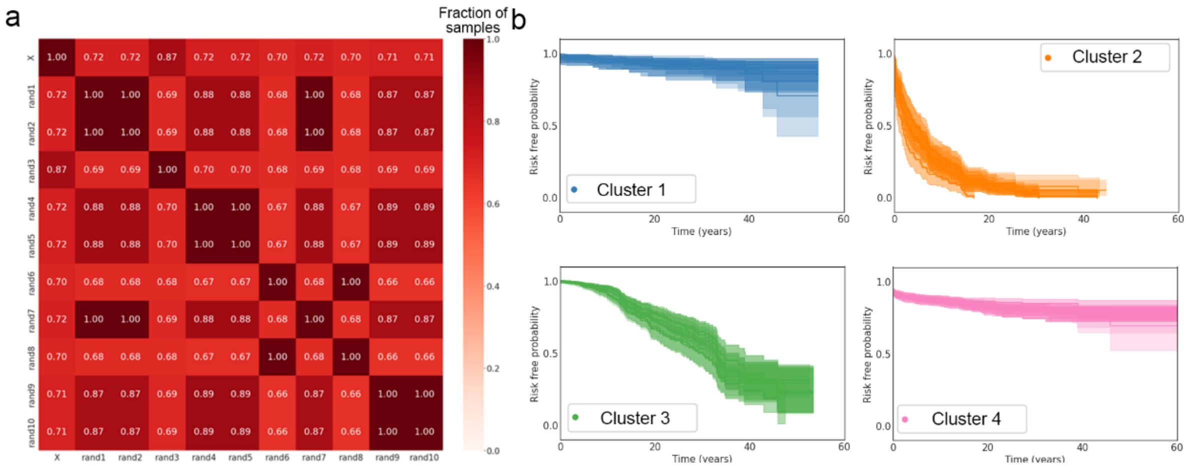


**Supplementary Fig. 7: Stability analysis for the CD application.** Results of running VaDeSCEHR 10 times with different initial weights and embeddings (rand1-10, n = 10). X indicates the result that we report as the main result in the manuscript. **a** Stability analysis for clustering. Each value indicates the fraction of the samples that obtain the same cluster assignment between two models. **b** Stability analysis for risk modeling. For each cluster, we plot the survival curve from 10 different models. Survival curves show a high degree of similarity across runs.

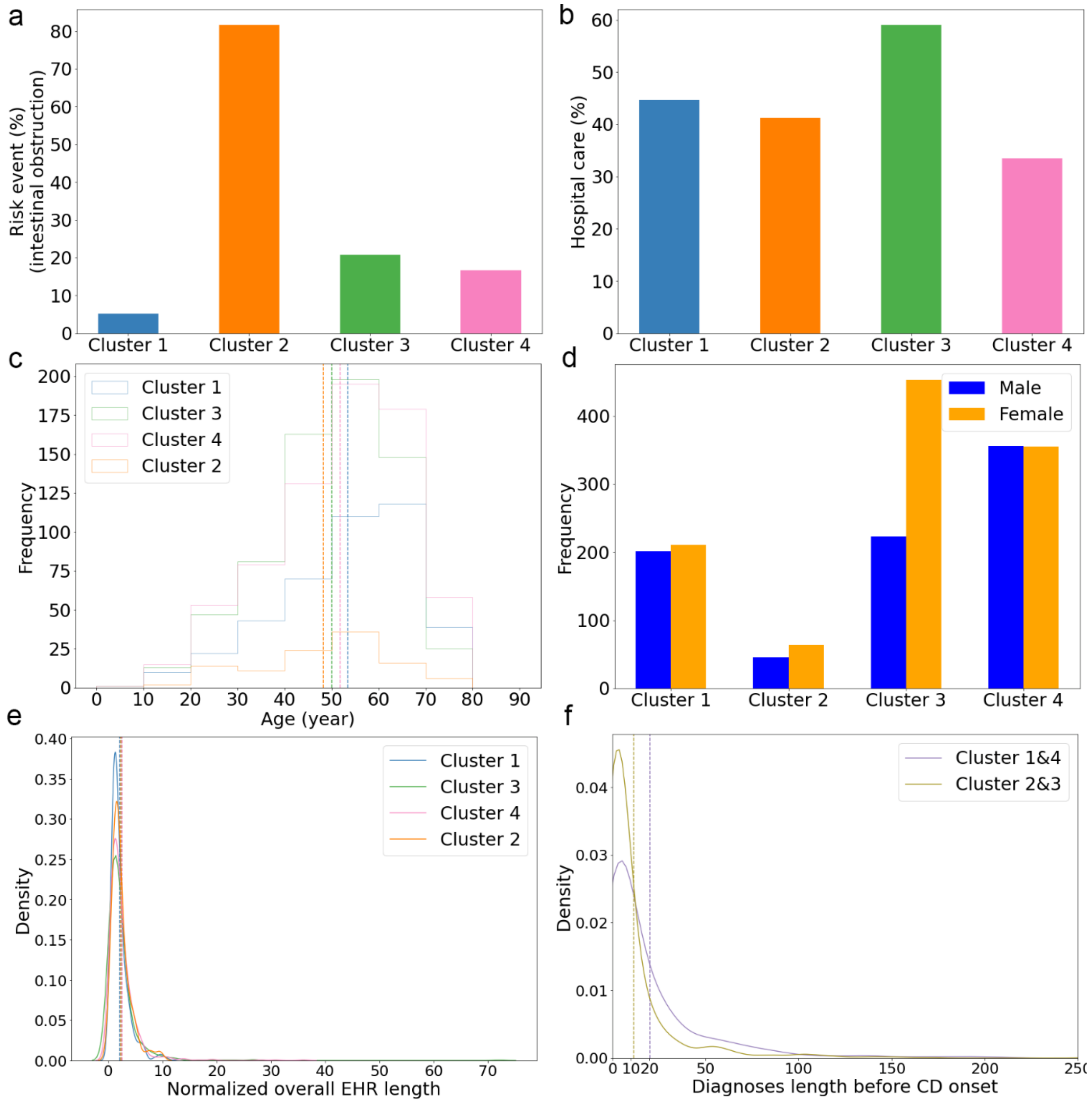


**Supplementary Fig. 8: Association of CD patient subgroups with different variables.** The analyses are based on 1,908 CD patients (n = 1,908). **a** Fraction of patients in each cluster with the risk event intestinal obstruction (p-value: $1.28 \times 10^{-50}$, two sided multinomial logistic regression with log likelihood ratio test, adjusting by confounding including age, sex, PC1, recruitment location, fraction of hospital care data). **b** Fraction of patients in each cluster with hospital care data (two sided Chi-squared p-value: $7.19 \times 10^{-20}$). **c** Distribution of age of onset in each cluster (two sided one-way ANOVA p-value: $6.82 \times 10^{-05}$). **d** Sex distribution in each cluster (two sided Chi-squared p-value: $2.35 \times 10^{-10}$). **e** Normalized overall diagnosis sequence length across clusters (two sided One-way ANOVA p-value: 0.19). **f** Distribution across clusters of diagnosis sequence length before first CD diagnosis (two sided one-way ANOVA p-value: $4.48 \times 10^{-10}$).

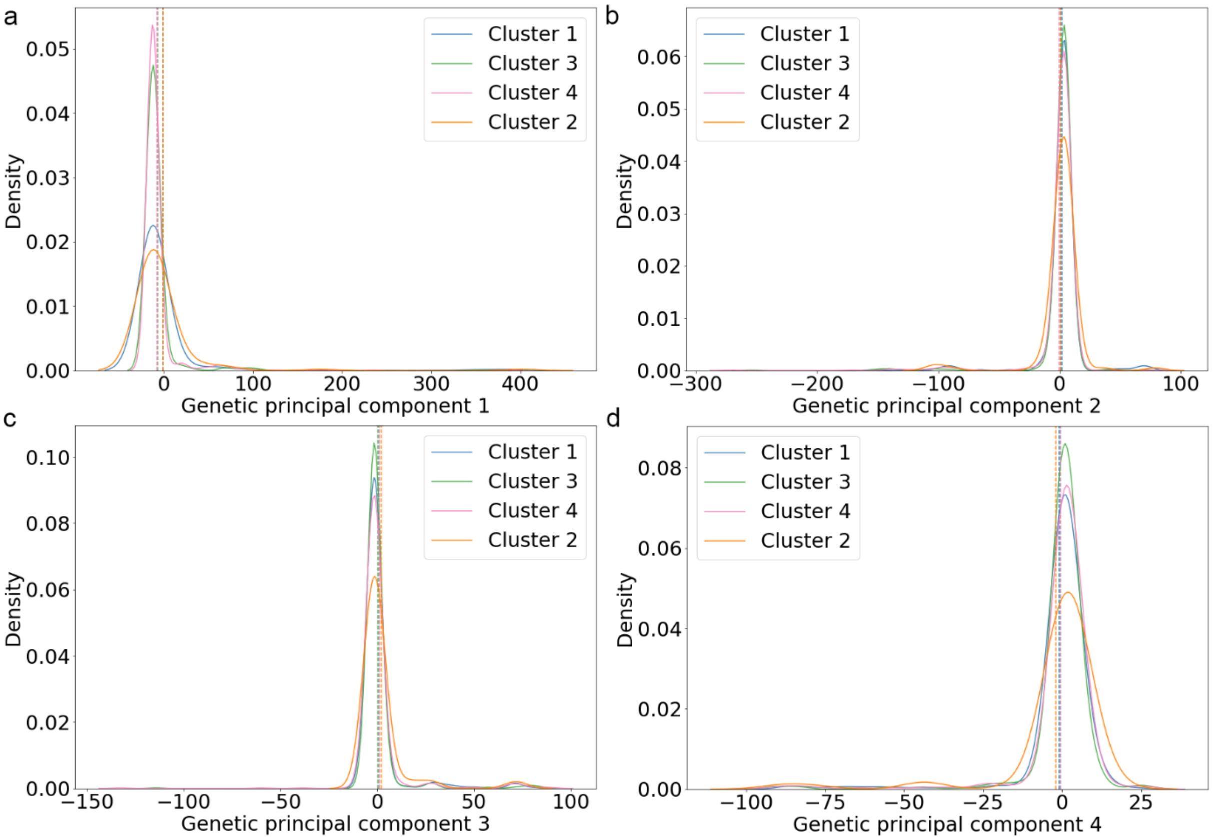


**Supplementary Fig. 9: Distribution of genetic principal components (PCs) across the four CD patient subgroups.** The analyses are based on 1,908 CD patients (n = 1,908). Two sided One-way ANOVA test was conducted to assess the significance of the difference between the clusters (p-value: 0.01, p-value: 0.29, p-value: 0.56, p-value: 0.64 for PC 1, 2, 3, and 4 respectively).

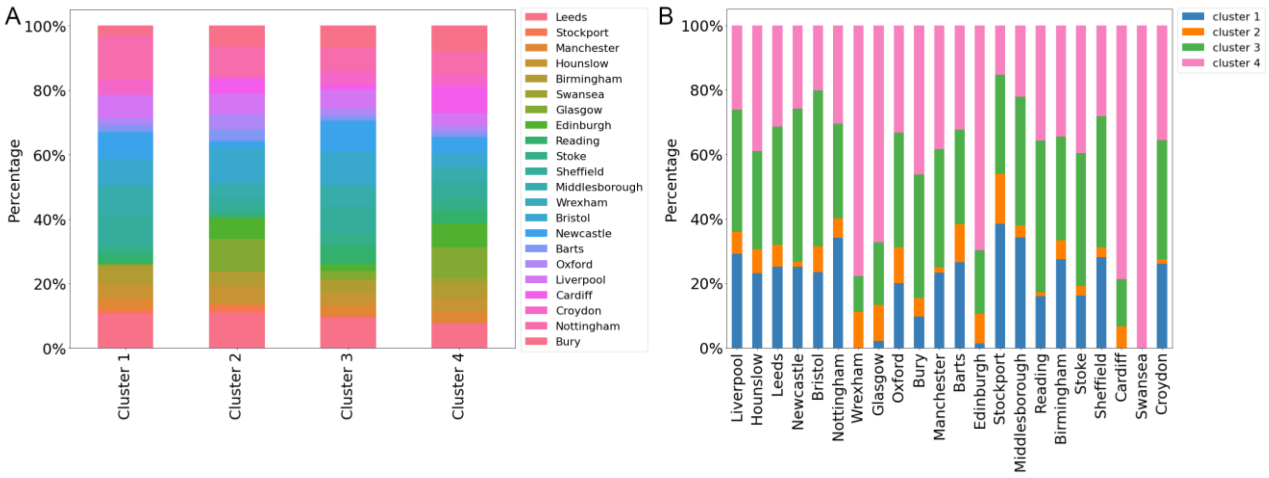


**Supplementary Fig. 10: Association of CD patient subgroups with location of UKB recruitment.** The analyses are based on 1,908 CD patients (n = 1,908) (permutation chi-squared p-val = 1). **a** proportion of locations in each cluster. **b** proportion of clusters in each location.

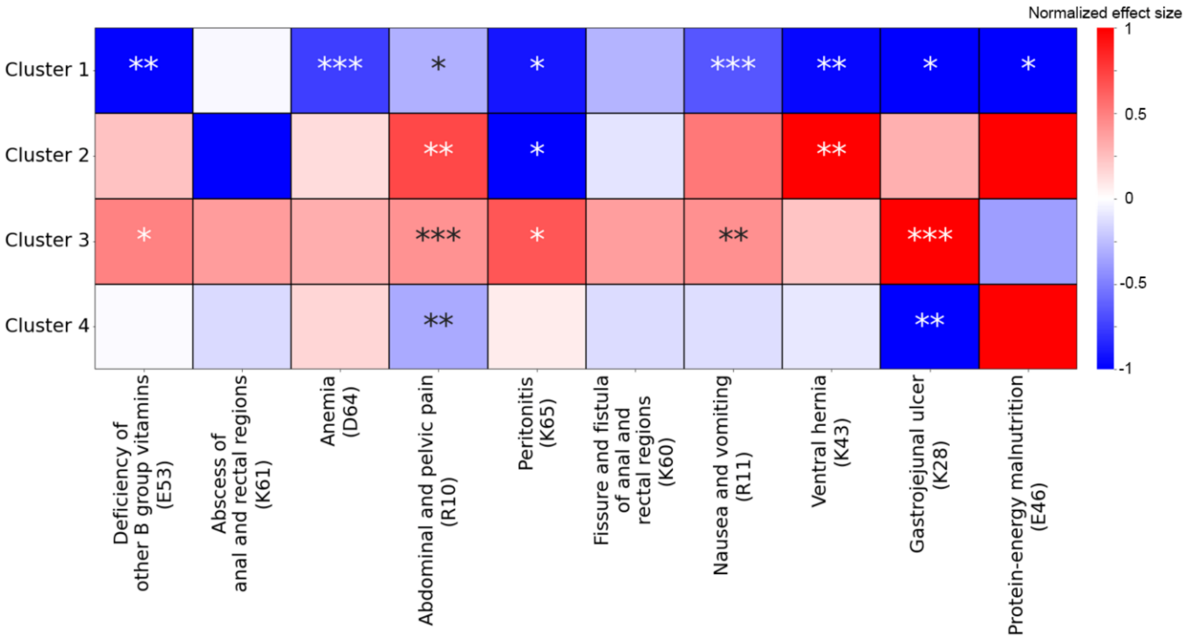


**Supplementary Fig. 11: Association of CD clusters with individual diagnoses.** The analyses are based on 1,908 CD patients (n = 1,908). Cluster-specific enrichment of individual CD-relevant diagnoses, computed relative to all other clusters, ranging from blue (negative) to red (positive) association. Asterisks indicate the significant level, adjusting p-values with the aim of controlling the false discovery rate (FDR) using the Benjamini-Hochberg procedure: * adjusted p-value < 0.05, ** adjusted p-value < 0.01 and *** adjusted p-value < 0.001.

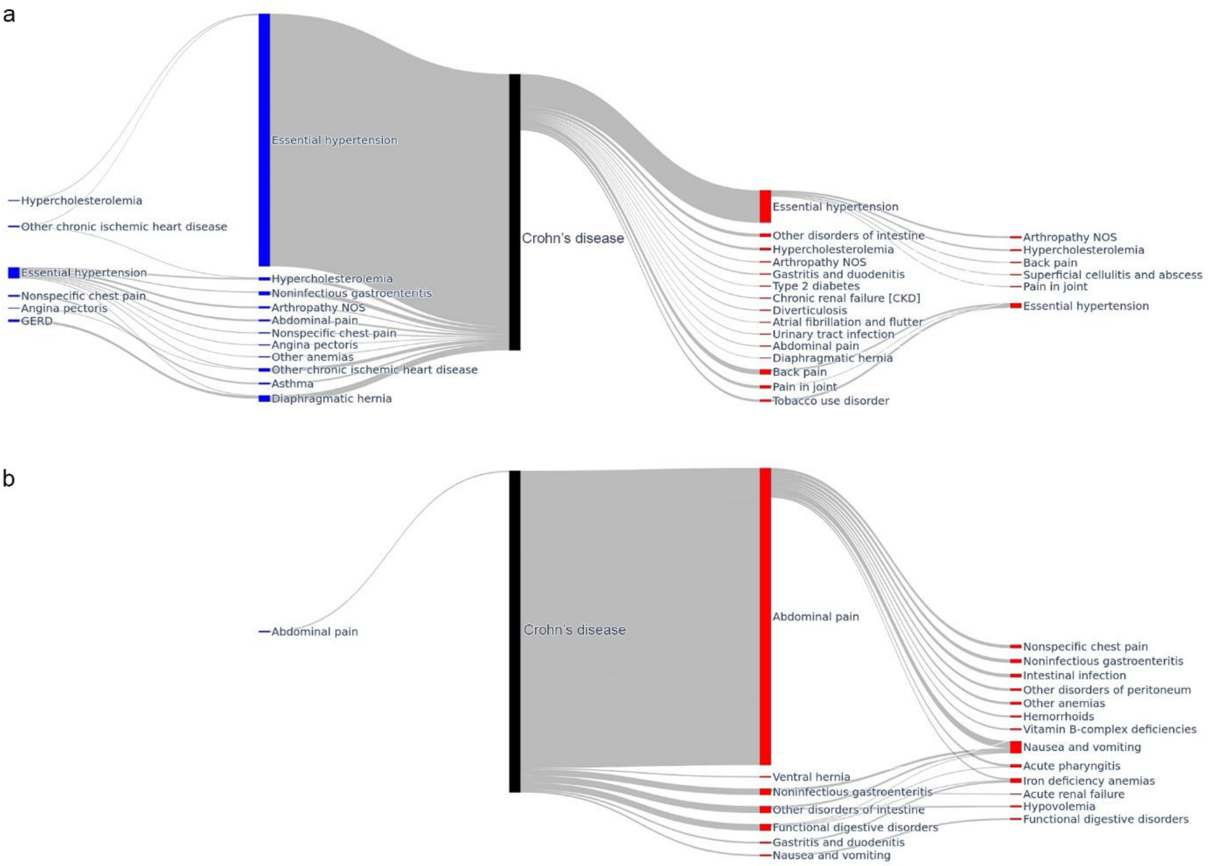


**Supplementary Fig. 12: Enrichment of diagnosis subsequences across fast and slowly progressing clusters.** Diagnoses are colored by occurrence before (blue) or after (red) the first CD diagnosis (black). **a** Diagnosis subsequences significantly enriched in slow progressors (clusters 1 and 4) relative to fast progressors (clusters 2 and 3) (logistic regression coefficient > 0 and adjusted p-value < 0.05) with the width of the band representing the number of patients with a given subsequence. **b** Diagnosis subsequences significantly enriched in fast progressors but not in slow progressors (logistic regression coefficient < 0 and adjusted p-value < 0.05).

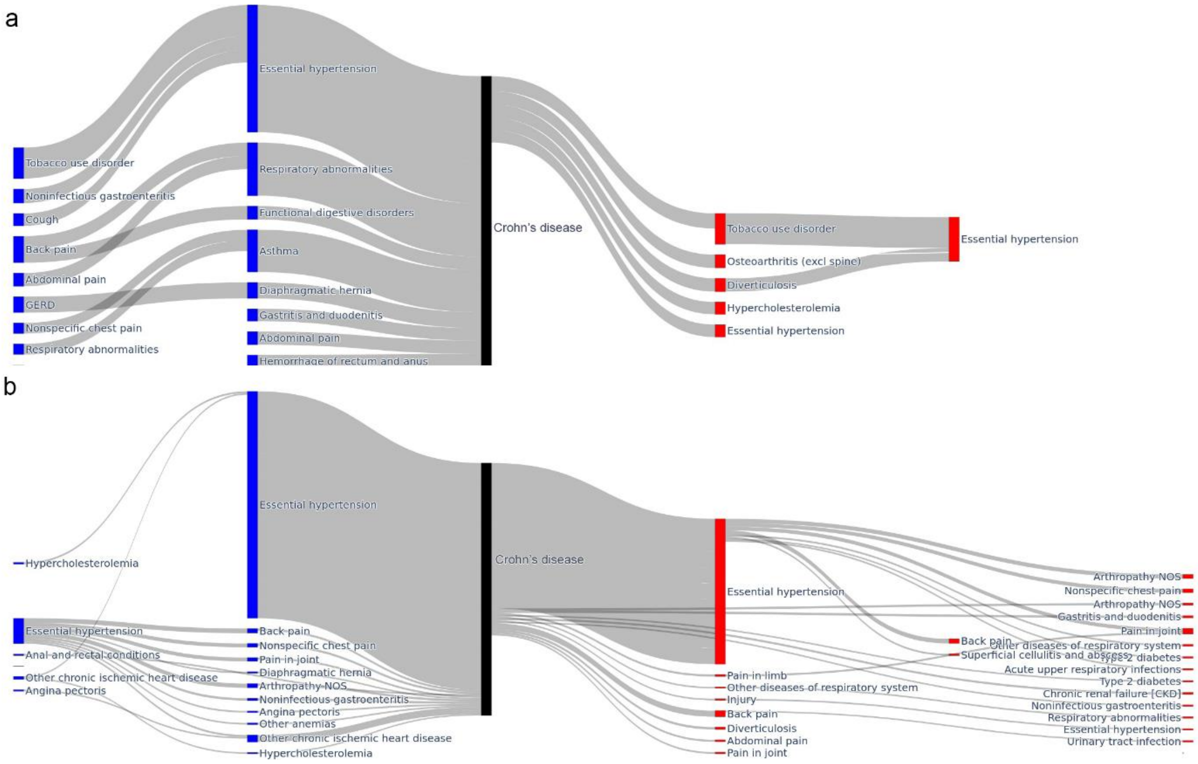


**Supplementary Fig. 13: Association of CD clusters with diagnosis subsequences.** Diagnosis subsequences enriched in cluster 1 **a** relative to cluster 4 **b** (logistic regression with a significance threshold at p-value: 0.05).

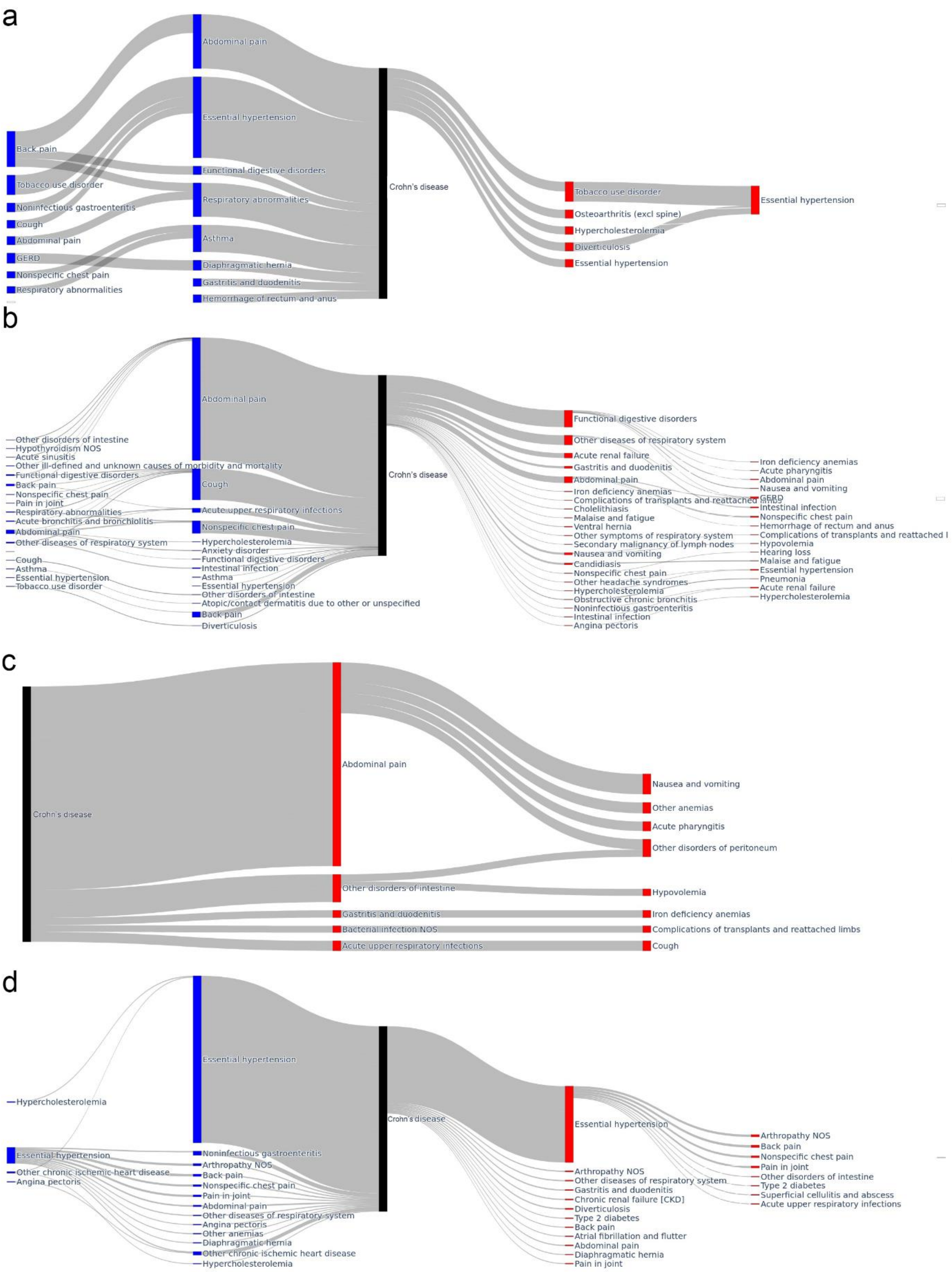


**Supplementary Fig. 14: Differentially enriched diagnosis subsequences per cluster. a** cluster 1, **b** cluster 2, **c** cluster 3, and **d** cluster 4. Statistical significance of association is determined using logistic regression (p-value < 0.05).

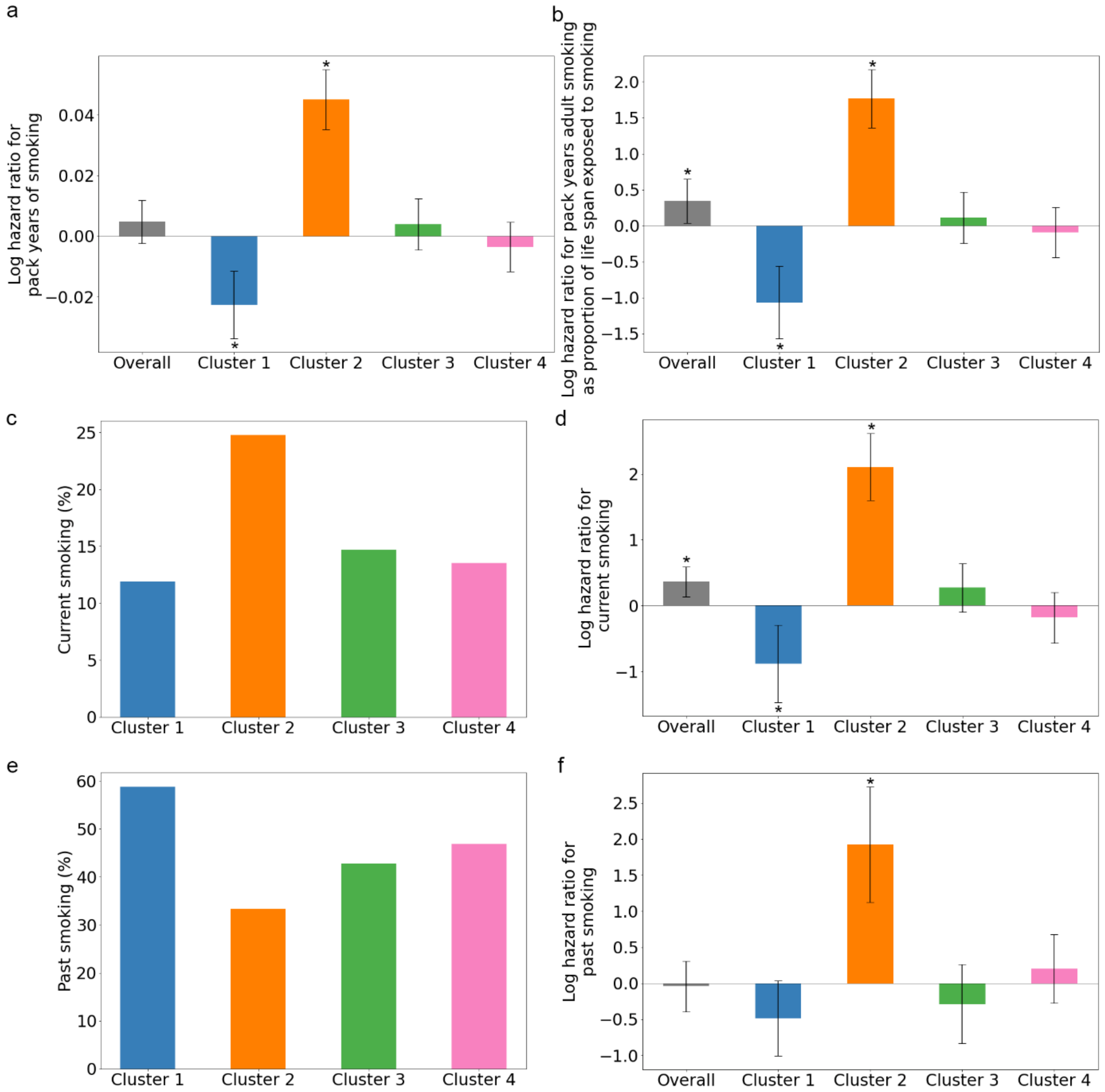


**Supplementary Fig. 15: Association of CD clusters with smoking behavior.** The analyses are based on 1,908 CD patients (n = 1,908). **a** Association of pack years of smoking (UK Biobank data field: 20161) with risk of intestinal obstruction (p-values are Overall: 0.19, Cluster 1: $7.17 \times 10^{-05}$, Cluster 2: $4.16 \times 10^{-19}$, Cluster 3: 0.37, Cluster 4: 0.39). **b** Association of pack years adult smoking as proportion of life span exposed to smoking with risk of intestinal obstruction (UK Biobank data field: 20162) (p-values are Overall: 0.03, Cluster 1: $3.40 \times 10^{-05}$, Cluster 2: $1.02 \times 10^{-17}$, Cluster 3: 0.55, Cluster 4: 0.59). **c** Percentage of current smokers (p-value: 0.038, multinomial logistic regression with log likelihood ratio test). **d** Association of current smoking with risk of intestinal obstruction (p-values are Overall: 0.002, Cluster 1: 0.003, Cluster 2: $6.56 \times 10^{-16}$, Cluster 3: 0.15, Cluster 4: 0.35). **e** Percentage previous smokers (p-value: $2.39 \times 10^{-9}$, multinomial logistic regression with log likelihood ratio test). **f** Association of past smoking with risk of intestinal obstruction (p-values are Overall: 0.81, Cluster 1: 0.07, Cluster 2: $2.56 \times 10^{-6}$, Cluster 3: 0.30, Cluster 4: 0.41). The error bars represent the 95% confidence intervals and * represents the significance of the association as determined using two sided multivariate Cox regression (p-value < 0.05).

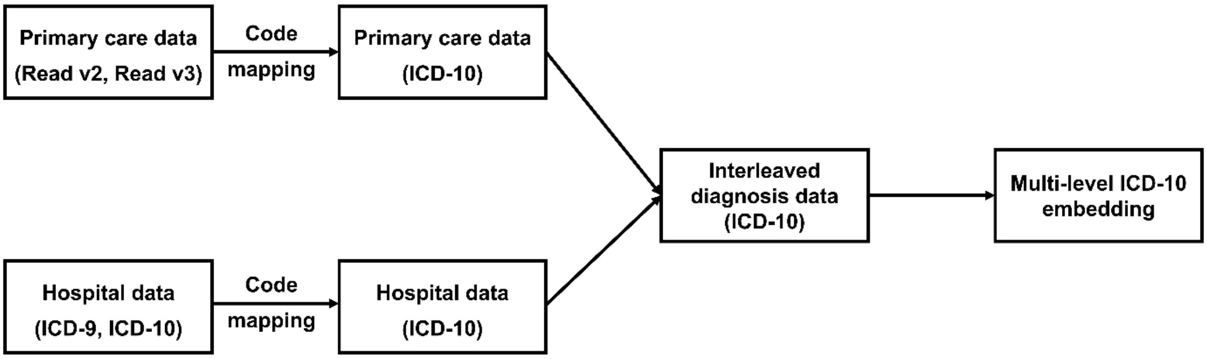


**Supplementary Fig. 16: UKB EHR data parser pipeline.** First, we map the various types of available diagnosis codes to ICD-10. Then we interleave the primary and hospital diagnoses based on their time stamps. Finally, we generate the multi-level ICD-10 embedding (details are described in the Method section).

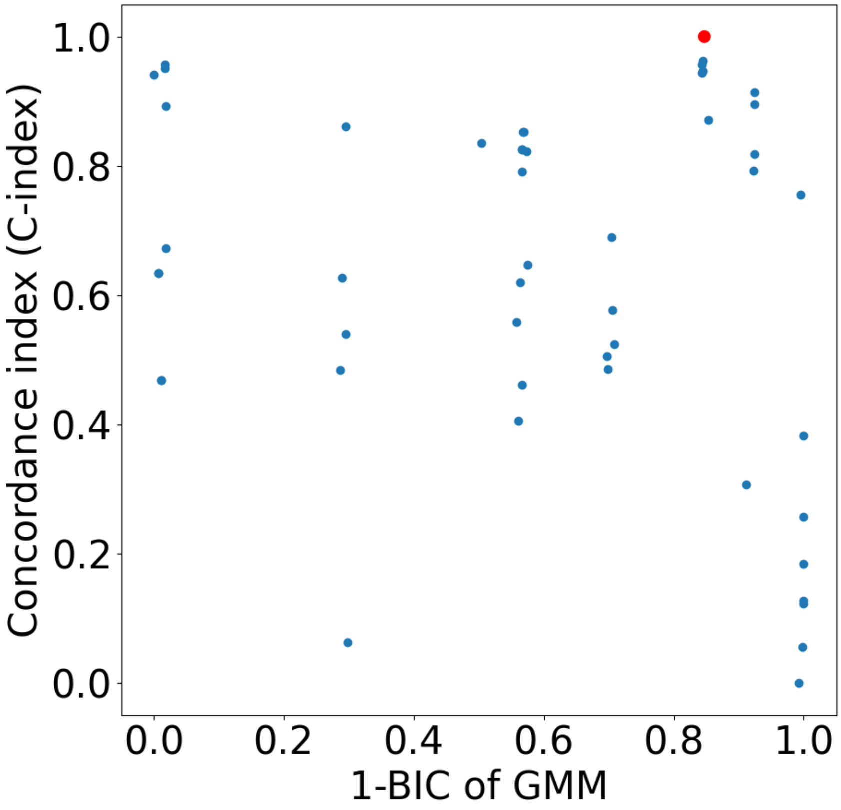


**Supplementary Fig. 17: Hyperparameter optimization for the CD application.** The number of clusters $\{1, 2, \dots, K\}$ was optimized jointly with the other hyperparameters. The best combination of hyperparameters (including the number of clusters) was determined by encouraging a low Bayesian information criterion (BIC) and a high concordance index (CI) through maximizing: $\sqrt{CI^2 + (1 - BIC_{norm})^2}$, where the BIC was normalized to the interval [0, 1]. The red dot represents the hyperparameter combination finally being selected with a cluster number of 4, which corresponds to the results of CD use case we describe in the main text. Details of the hyperparameters are shown in Supplementary Table 4.

**Supplementary Table 1: Summary of the data sets used in this study (supplementing Table 1)**

| | | UK Biobank EHR | T1D/T2D benchmark | CD application |
|---|---|---|---|---|
| **Location** | Glasgow | 18644 | 98 | 41 |
| | Middlesborough | 21283 | 108 | 106 |
| | Cardiff | 17875 | 75 | 67 |
| | Leeds | 44185 | 175 | 158 |
| | Bristol | 43000 | 149 | 213 |
| | Newcastle | 36995 | 135 | 221 |
| | Nottingham | 33870 | 167 | 122 |
| | Bury | 28311 | 123 | 134 |
| | Reading | 29399 | 81 | 90 |
| | Birmingham | 25493 | 84 | 121 |
| | Liverpool | 32799 | 103 | 197 |
| | Manchester | 13937 | 60 | 108 |
| | Sheffield | 30381 | 135 | 116 |
| | Barts | 12574 | 34 | 63 |
| | Oxford | 14054 | 45 | 58 |
| | Croydon | 27363 | 73 | 182 |
| | Hounslow | 28866 | 82 | 172 |
| | Edinburgh | 17193 | 76 | 33 |
| | Stoke | 19426 | 68 | 88 |
| | Stockport | 3791 | 13 | 23 |
| | Swansea | 2280 | 15 | 6 |
| | Glasgow | 18644 | 98 | 41 |

**Supplementary Table 2: Trials run as part of Bayesian hyperparameter optimization for the pretraining of transformer encoder.**

| trials | hidden_size | num_attention_h | num_hidden_laye | intermediate_size | initializer_range | dropout | learning_rate | weight_decay | loss |
|---|---|---|---|---|---|---|---|---|---|
| 1 | 384 | 12 | 9 | 2048 | 0.05 | 0.2 | 0.001 | 1.00E-06 | 1.33867 |
| 2 | 384 | 8 | 12 | 1024 | 0.04 | 0.2 | 1.00E-05 | 0.0001 | 0.99953 |
| 3 | 768 | 12 | 9 | 1024 | 0.02 | 0.3 | 1.00E-06 | 1.00E-05 | 1.21922 |
| 4 | 768 | 16 | 9 | 1024 | 0.02 | 0.1 | 0.0001 | 0.0001 | 0.54278 |
| 5 | 768 | 16 | 12 | 1280 | 0.01 | 0.5 | 0.0001 | 0.01 | 0.89176 |
| 6 | 384 | 12 | 9 | 1280 | 0.04 | 0.1 | 0.001 | 1.00E-06 | 1.10544 |
| 7 | 384 | 16 | 12 | 2048 | 0.02 | 0.1 | 1.00E-05 | 0.0001 | 0.87461 |
| 8 | 384 | 8 | 3 | 1024 | 0.04 | 0.1 | 0.001 | 0.1 | 0.85247 |
| 9 | 768 | 12 | 6 | 1280 | 0.02 | 0.2 | 0.0001 | 0.1 | 0.64979 |
| 10 | 384 | 16 | 9 | 1024 | 0.02 | 0.2 | 1.00E-05 | 1.00E-05 | 0.99285 |
| 11 | 768 | 8 | 6 | 1024 | 0.02 | 0.2 | 0.0001 | 0.0001 | 0.68340 |
| 12 | 384 | 16 | 12 | 1024 | 0.05 | 0.3 | 0.0001 | 1.00E-05 | 0.83419 |
| 13 | 768 | 16 | 6 | 1280 | 0.03 | 0.2 | 1.00E-05 | 0.0001 | 1.00722 |
| 14 | 384 | 12 | 9 | 1024 | 0.01 | 0.1 | 1.00E-06 | 0.001 | 1.04359 |
| 15 | 768 | 16 | 3 | 2048 | 0.01 | 0.1 | 1.00E-06 | 0.001 | 1.21871 |
| 16 | 768 | 12 | 3 | 1280 | 0.01 | 0.3 | 0.0001 | 0.001 | 0.48211 |
| **17** | **768** | **16** | **6** | **1280** | **0.02** | **0.2** | **0.0001** | **0.001** | **0.41522** |
| 18 | 768 | 12 | 6 | 1280 | 0.03 | 0.1 | 0.0001 | 0.1 | 0.41944 |
| 19 | 768 | 16 | 3 | 2048 | 0.03 | 0.3 | 0.0001 | 0.01 | 0.48143 |
| 20 | 768 | 8 | 3 | 2048 | 0.03 | 0.3 | 0.0001 | 0.01 | 0.46234 |

**Supplementary Table 3: Trials run as part of Bayesian hyperparameter optimization for the fine-tuning of CD use case.**

| trials | num_cluster | dropout_ prob | epoch | latent_dim | learning_ rate | num_reshape _layers | weibull _shape | weight_decay | Value |
|---|---|---|---|---|---|---|---|---|---|
| 1 | 2 | 0.4 | 50 | 5 | 1.00E-05 | 2 | 1 | 0.001 | 1.007 |
| 2 | 2 | 0.4 | 50 | 5 | 0.001 | 2 | 1 | 0.001 | 1.016 |
| 3 | 2 | 0.3 | 50 | 20 | 5.00E-05 | 4 | 4 | 0.001 | 0.692 |
| 4 | 2 | 0.1 | 100 | 5 | 1.00E-05 | 2 | 1 | 0.001 | 1.007 |
| 5 | 2 | 0.1 | 100 | 5 | 0.001 | 2 | 1 | 0.001 | 1.070 |
| 6 | 2 | 0.3 | 100 | 5 | 0.001 | 4 | 1 | 0.001 | 1.032 |
| 7 | 2 | 0.5 | 100 | 20 | 0.001 | 1 | 3 | 0.0001 | 0.789 |
| 8 | 2 | 0.2 | 150 | 5 | 1.00E-05 | 2 | 1 | 0.0001 | 1.000 |
| 9 | 2 | 0.2 | 150 | 10 | 0.0005 | 3 | 3 | 1.00E-05 | 1.220 |
| 10 | 2 | 0.5 | 150 | 15 | 5.00E-05 | 1 | 3 | 0.0001 | 0.881 |
| 11 | 2 | 0.2 | 150 | 15 | 0.0005 | 4 | 4 | 1.00E-05 | 0.911 |
| 12 | 2 | 0.1 | 150 | 20 | 0.001 | 3 | 3 | 0.0001 | 0.973 |
| 13 | 2 | 0.3 | 200 | 15 | 0.0001 | 4 | 3 | 1.00E-05 | 0.861 |
| 14 | 2 | 0.2 | 200 | 20 | 1.00E-05 | 2 | 2 | 0.01 | 0.731 |
| 15 | 2 | 0.3 | 200 | 20 | 0.0005 | 4 | 3 | 1.00E-05 | 0.837 |
| 16 | 2 | 0.4 | 300 | 5 | 1.00E-05 | 2 | 1 | 1.00E-06 | 0.992 |
| 17 | 2 | 0.4 | 300 | 5 | 0.001 | 3 | 5 | 1.00E-06 | 1.249 |
| 18 | 2 | 0.4 | 300 | 15 | 0.0001 | 4 | 3 | 1.00E-05 | 0.850 |
| 19 | 3 | 0.4 | 50 | 5 | 0.0001 | 4 | 5 | 1.00E-05 | 1.287 |
| 20 | 3 | 0.1 | 50 | 20 | 0.001 | 1 | 3 | 0.0001 | 0.690 |
| 21 | 3 | 0.1 | 50 | 20 | 0.001 | 2 | 3 | 0.0001 | 0.700 |
| 22 | 3 | 0.4 | 100 | 5 | 0.0001 | 4 | 5 | 1.00E-05 | 1.288 |
| 23 | 3 | 0.3 | 150 | 5 | 1.00E-05 | 3 | 4 | 0.0001 | 1.217 |
| 24 | 3 | 0.3 | 150 | 5 | 1.00E-05 | 4 | 4 | 0.0001 | 1.219 |
| 25 | 3 | 0.1 | 150 | 15 | 1.00E-05 | 1 | 4 | 0.01 | 0.976 |

| | | | | | | | | | |
|---|---|---|---|---|---|---|---|---|---|
| 26 | 3 | 0.2 | 200 | 5 | 0.0005 | 3 | 2 | 0.001 | 1.234 |
| 27 | 3 | 0.1 | 200 | 5 | 0.0005 | 4 | 3 | 0.001 | 1.300 |
| 28 | 3 | 0.4 | 300 | 10 | 0.0001 | 2 | 3 | 0.001 | 0.986 |
| 29 | 3 | 0.3 | 300 | 20 | 0.0001 | 4 | 2 | 1.00E-06 | 0.563 |
| 30 | 3 | 0.4 | 300 | 20 | 0.0001 | 4 | 2 | 1.00E-06 | 0.573 |
| 31 | 4 | 0.3 | 50 | 10 | 0.0001 | 3 | 4 | 0.0001 | 1.002 |
| 32 | 4 | 0.5 | 100 | 10 | 1.00E-05 | 1 | 2 | 0.0001 | 0.865 |
| 33 | 4 | 0.4 | 100 | 10 | 0.0001 | 2 | 4 | 1.00E-05 | 1.025 |
| 34 | 4 | 0.5 | 100 | 15 | 1.00E-05 | 1 | 1 | 1.00E-06 | 0.616 |
| 35 | 4 | 0.1 | 100 | 20 | 5.00E-05 | 2 | 1 | 0.01 | 0.468 |
| 36 | 4 | 0.2 | 100 | 20 | 1.00E-05 | 1 | 2 | 1.00E-06 | 0.673 |
| 37 | 4 | 0.5 | 100 | 20 | 0.0005 | 1 | 2 | 0.01 | 0.634 |
| 38 | 4 | 0.3 | 150 | 5 | 5.00E-05 | 4 | 5 | 1.00E-05 | 1.281 |
| 39 | 4 | 0.4 | 150 | 10 | 0.0001 | 2 | 4 | 0.0001 | 0.962 |
| 40 | 4 | 0.3 | 200 | 5 | 5.00E-05 | 4 | 5 | 1.00E-05 | 1.275 |
| 41 | 4 | 0.5 | 200 | 5 | 5.00E-05 | 4 | 5 | 0.0001 | 1.268 |
| 42 | 4 | 0.4 | 200 | 10 | 5.00E-05 | 3 | 3 | 1.00E-05 | 1.001 |
| 43 | 4 | 0.1 | 200 | 20 | 1.00E-05 | 1 | 3 | 0.01 | 0.956 |
| 44 | 4 | 0.3 | 250 | 5 | 5.00E-05 | 4 | 5 | 1.00E-05 | 1.266 |
| 45 | 4 | 0.5 | 250 | 15 | 1.00E-05 | 1 | 5 | 1.00E-06 | 0.911 |
| 46 | 4 | 0.1 | 250 | 20 | 1.00E-05 | 4 | 4 | 0.01 | 0.893 |
| **47** | **4** | **0.4** | **300** | **5** | **0.0005** | **2** | **5** | **1.00E-06** | **1.311** |
| 48 | 4 | 0.1 | 300 | 15 | 0.001 | 2 | 1 | 1.00E-05 | 0.305 |
| 49 | 4 | 0.4 | 300 | 20 | 1.00E-05 | 1 | 2 | 0.001 | 0.952 |
| 50 | 4 | 0.1 | 300 | 20 | 5.00E-05 | 1 | 5 | 1.00E-06 | 0.941 |
