## Supplementary Data 1(description) for "Deep representation learning for clustering longitudinal survival data from electronic health records"

**Supplementary Data 1: Results of pathway PRS.** Significance assessed using logistic regression with log likelihood ratio test, which was a two-sided test. And multiple testing corrected using the Benjamini–Hochberg procedure.
